# Community prevalence of antibodies to SARS-CoV-2 and correlates of protective immunity in an Indian metropolitan city

**DOI:** 10.1101/2020.11.17.20228155

**Authors:** Aurnab Ghose, Sankar Bhattacharya, Arun S. Karthikeyan, Abhay Kudale, Joy M. Monteiro, Aparna Joshi, Guruprasad Medigeshi, Gagandeep Kang, Vineeta Bal, Satyajit Rath, L. S. Shashidhara, Jacob John, Susmita Chaudhuri, Aarti Nagarkar

## Abstract

**Objectives:** To assess seroprevalence of anti-SARS-CoV-2 antibodies in a densely populated urban Indian settings and its implications for disease trends and protective immunity.

**Design:** Cross-sectional sero-epidemiological survey linked with administrative reporting of COVID-19 testing data.

**Settings:** Pune city in western India

**Main outcome measure:** Prevalence of anti-SARS-CoV-2 spike protein antibodies were estimated and along with correlates of virus neutralisation and other immune and inflammatory markers.

**Results:** Seropositivity was extensive (51·3%; 95%CI 39·9 to 62·4) but varied widely in the five localities tested, ranging from 35·8% to 66·4%. Seropositivity was higher in crowded living conditions in the slums (OR 1·91), and was lower in those 65 years or older (OR 0·59). The infection-fatality ratio was estimated at 0.21%. Post survey, COVID-19 incidence was lower in areas noted to have higher seroprevalence. Substantial virus-neutralising activity was observed in seropositive individuals, but with considerable heterogeneity in the immune response and possible age-dependent diversity in the antibody repertoire.

**Conclusion:** Despite crowded living conditions having facilitated widespread transmission, the variability in seroprevalence in localities that are in geographical proximity indicates a heterogenous spread of infection. Declining infection rates in areas with high seropositivity suggest population-level protection. It is also supported by substantial neutralising activity in asymptomatically infected individuals. This is the first report of a significantly high proportion of protective immune response among asymptomatic individuals in the population. The heterogeneity in antibody levels and neutralisation capacity indicates the existence of immunological sub-groups of functional interest.

**Trial registration:** Registered with the Clinical Trials Registry of India (CTRI/2020/07/026509)

## INTRODUCTION

Examining the seroprevalence of Severe Acute Respiratory Syndrome Coronavirus 2 (SARS-CoV-2)-specific antibodies in areas with significant spread of the virus is important for estimating the proportion of the population infected by the virus. In addition, seroprevalence studies can clarify the spread of asymptomatic infection and provide insights into the extent to which apparently low infection (and case) fatality rates have been confounded by prevalent testing strategies. High seropositivity in communities may modify transmission rates through population-level protection. The survey of literature (up to 2 Nov 2020) indicated that community serosurveys for SARS-CoV-2 in LICs and LMICs have been limited and have largely reported correlations of seroprevalence with demographic factors. Only a few studies in the general population report double-digit seropositivity. Of these, most are from high-income/upper-middle-income countries^1 2^ with only a few from low-income and lower-middle-income countries (LICs/LMICs)^1 3^. Notably, the latter studies have reported very high seroprevalence^1 3^. There are no reports of protective immunity-associated characteristics in community surveillance settings from LMIC/LICs. In fact, such studies from the global North are also limited. The existing evidence thus lacks granular details critical to understand community-level heterogeneities, and provides limited epidemiological data without meaningful immunobiological correlates.

The spread of COVID-19 is likely to be affected by local demographics, socio-economic conditions, the history and burden of infectious diseases as well as biological factors but there are very few studies from LMICs with sufficient local granularity and none explore disease prevalence vis-à-vis correlates of protective immunity.

Pune, a metropolitan city in Western India, recorded its first confirmed case of COVID-19 on March 9, 2020 and by November 24, 2020 has reported 167,604 cases and 4,440 deaths. We undertook a cross-sectional sero-epidemiological study in five high-incidence administrative subwards of Pune to estimate the cumulative burden of the disease in these communities and to examine the associated risk-factors. As antibodies against the receptor-binding domain (RBD) of the viral spike protein are predominantly associated with neutralising activity, we estimated the seroprevalence of anti-RBD IgG (RBD-IgG) antibodies^4 5^.

Given the high proportion of asymptomatic SARS-CoV-2 infections^6^, investigating the putative correlates of immune protection in such individuals is necessary to understand the basis of community-level protection. The immune response to SARS-CoV-2 infection is expected to be heterogeneous within populations and an understanding of this diversity is likely to be central to the management of the disease. We have therefore characterised the RBD-specific antibody responses for their ability to prevent cellular receptor binding, as well as for virus neutralisation. Finally, elevated markers of inflammation have been associated with poor prognosis in clinical cases of COVID-19^7 8^, and unresolved systemic inflammation is increasingly being thought to underlie some of the heterogeneous, long-term sequelae of COVID-19^9-13^. However, it is not clear if inflammation is associated with asymptomatic infections. Thus, we have also tested a parameter of systemic inflammation in the sampled population.

## METHODS

### Study design, setting and analysis of seroprevalence

The PMC administrative area is divided into 41 subwards with an average population of 76,394 per subward. Based on the incidence of RT-PCR confirmed COVID-19 cases as on 31 May 2020, we stratified the sub-wards as low (<0.62 per 1000), medium (0.61 to 2.5 per 1000), and high incidence (>2.5 per 1000) settings. Five subwards were randomly selected from among the 13 subwards (∼400,000 population) classified as high incidence settings for a sero-epidemiological survey. The sampling strategy is summarized as a flow chart in Figure 1. An independent team of geospatial experts divided the 5 selected subwards into 235 polygonal grids of roughly equal area and randomly selected 63 of the 235 grids for sampling (12-14 grids per subward). From the approximate center of each grid, participants were recruited from every fifth house with sequential recruitment in multi-occupancy tenements as well. One consenting adult per household, above 18 years of age was selected following a pre-decided age and sex distribution criteria^14^.

**Figure 1:**
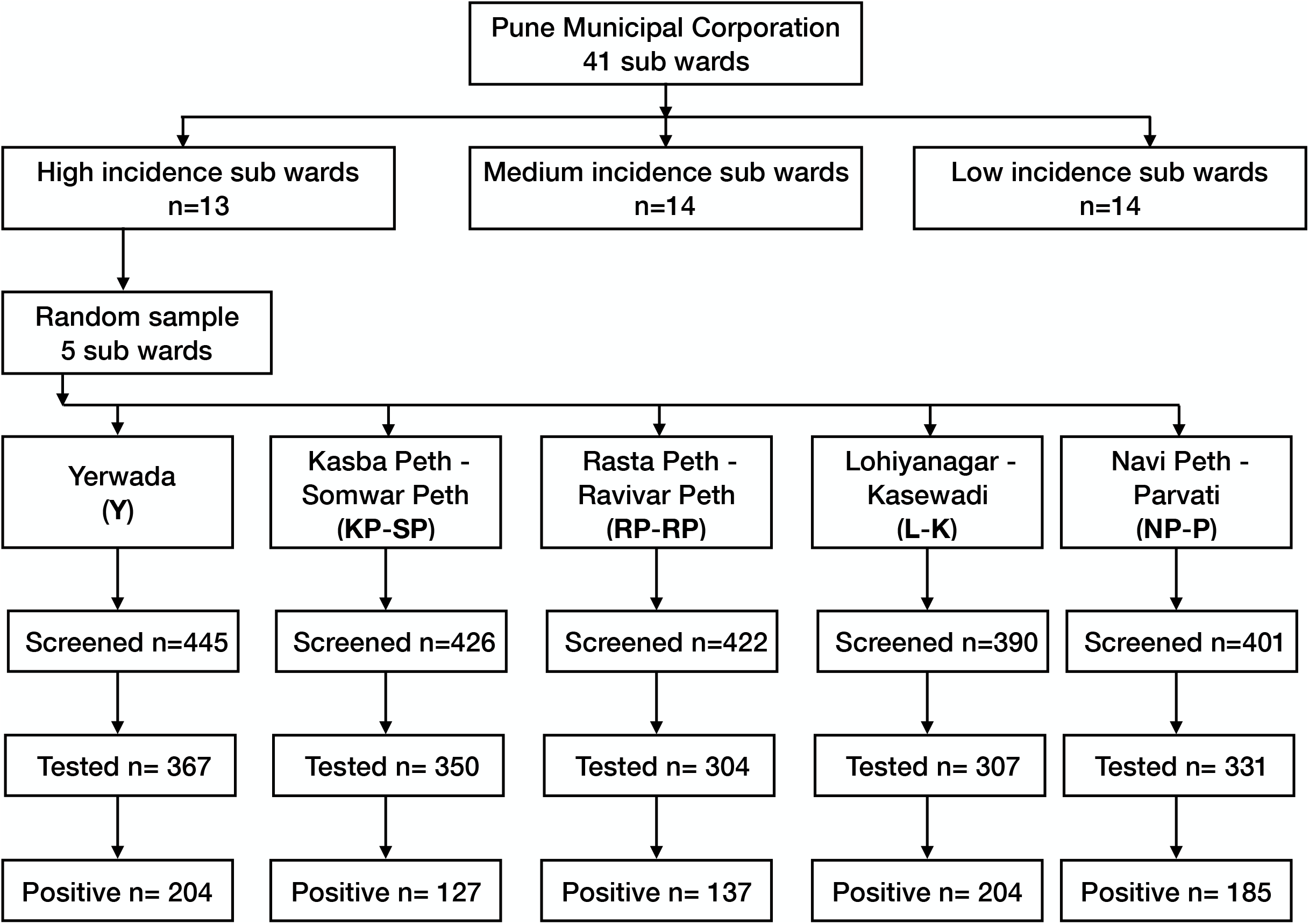
Flowchart summarising the sampling strategy and seropositivity.

Active containment zones were excluded because of operational challenges. Currently ill, febrile individuals were excluded, though, those who had an illness episode, including COVID-19, in the past but were well at recruitment remained eligible for recruitment.

The study team visited selected households and after written informed consent, five ml of peripheral venous blood was collected. Blood samples were stored in icepacks and transported to the laboratory for processing within 5 hours of collection.

#### Study subjects and sample size

Considering a possible 15% serological prevalence in the selected clusters, for a relative precision of 20%, a confidence interval of 95%, and with a design effect of 2.5, 1360 participants would be required. The sample size was inflated by 20% to account for inadequate samples and uniform recruitments from each cluster.

### Serum isolation and immunoassays

Serum was separated using standard methods and stored in multiple aliquots. The in-house THSTI RBD-IgG ELISA was performed using methods previously described^15^. We assumed that the IgG antibodies are detectable 14 to 21 days post-infection and that these antibodies persist, following infection, for four months^4 16 17^. No adjustments were made for assay accuracy and the potential impact of declining antibodies on seroprevalence.

ELISA-based surrogate virus neutralisation test (sVNT; Genscript), SARS-CoV-2 S1 IgA ELISA (Eurimmune) and high-sensitivity C-reactive protein ELISA (Diagnostic Biochem) were performed according to manufacturer’s protocols.

SARS-CoV-2 (isolate USA-WA1/2020) virus expanded in Vero E6 cells was used with heat-inactivated sera for Plaque Reduction Neutralization Tests (PRNTs). The PRNT_50_ neutralisation titres are the inverse endpoint dilutions of sera that could neutralize 50% of viral plaque formation.

### Statistical methods and presentation of results

The analysis accounted for the multistage clustered survey methods by assigning sampling weightage to adjust for the probability of selection of each sampling unit. Logistic regression with linearized standard errors was used for examining factors associated with being seropositive. The reproduction numbers at different time points were estimated for each sub ward using EpiEstim package^18^. To estimate the association between seroprevalence and incidence of new cases post survey we performed a linear regression for the incidence of new cases, adjusting for the reported incidence at the beginning of the survey. We did not adjust prevalence for imperfect test sensitivity, since there is currently no gold standard for comparison and as the sensitivity of the assay used compared favourably with other assays^15^.

We estimated IFR as the ratio of the number of reported COVID-19 deaths by 23 July 2020 to the estimated number of infected individuals in the population. We assumed a 13 day median interval between infection and mortality among those who died^19^ and that seroprevalence reflects incidence two weeks prior to survey.

#### Variables, data sources, and bias

We obtained demographic and household characteristics and collected information on potential risk factors including occupation, exposure to SARS CoV-2 including travel history during the lockdown period, experience of symptoms and history of quarantine. These data were collected independent of serology using a standardized questionnaire at the initial survey.

#### Statistical tests and software

Analyses were performed using STATA v14.2 13.1, R Statistical Software v2.14.0 and GraphPad Prism v8.1.2. Spearman’s non-parametric correlation analysis was done for dual datasets. Mann-Whitney test or Kruskal-Wallis ANOVA with Dunn’s post-test was used to compare across groups. SPSS was used for regression analysis of sex and age data with serological tests and categorical comparisons made using the Chi-square test.

Box-and-whisker plots: the box extends from the 25th to 75th percentiles with the median indicated by a line. The whiskers mark the 5th and 95th percentiles.

### Ethical considerations

The study was approved by the Institutional Ethics Committee of SPPU (SPPU/IEC/2020/82), IISER Pune (IECHR/Admin/2020/005) and THSTI (THS1.8.1(106)) and the Institutional Biosafety Committees of IISER Pune and THSTI. The study was prospectively registered with the Clinical Trials Registry of India (CTRI/2020/07/026509).

## RESULTS

### Seroprevalence of anti-SARS-CoV-2 IgG antibodies

Between 20^th^ July and 5^th^ August 2020,1659 eligible, consenting adult participants were recruited from 2089 screened individuals (Figure 1). The study recruited 801 women and 858 men from the five subwards with demographic characteristics as shown in Table 1. 857 individuals tested positive for the anti-RBD IgG ELISA. The sample weighted seroprevalence was estimated at 51·3% (95%CI 39·9 to 62·4).

**Table 1:** Demographic characteristics of the study population

| Characteristics | N (%) |
| --- | --- |
| **N** | 1659 |
| **Subward** |  |
| Yerwada | 367 (22·1) |
| Kasba Peth-Somwar Peth | 350 (21·1) |
| Rasta Peth-Ravivar Peth | 304 (18·3) |
| Lohiyanagar-Kasewadi | 307 (18·5) |
| Navi Peth-Parvati | 331 (20·0) |
| **Age categories [years]** |  |
| 18-30 | 394 (23·8) |
| 31-50 | 677 (40·8) |
| 51-65 | 417 (25·1) |
| 66-above | 171 (10·3) |
| **Gender** |  |
| Male | 801 (48·3) |
| Female | 858 (51·7) |
| **Family size** (mean (SD)) | 4·6 (2·3) |
| **Education in years** (mean (SD)) | 9·14 (5·9) |
| **Type of dwelling** |  |
| Non-slums | 527 (31·8) |
| Slums | 1132 (68·2) |
| **Floor area per person [m<sup>2</sup>]**<br>(median (IQR)) | 7·0 (4·3-11·1) |
| **Shared toilet** | 616 (37·1) |
| **Contact with case** | 103 (6·2) |
| **History of symptoms** | 105 (6·3) |
| **History of COVID-19** | 21 (1·3) |

Notably, there was considerable heterogeneity in seroprevalence between clusters (Table 2; Figure 2). Kasbapeth-Somvarpeth subward had the lowest seroprevalence at 35·8% (95%CI 29·2 to 43·1) while the Lohiyanagar subward had the highest seroprevalence of 66·4% (95%CI 57·8 to 74·1). The differences in seroprevalence by characteristics of the study population are presented in Table 2. The seroprevalence across age-groups was uniform in those under 66 years of age. The oldest age-group had a significantly lower seroprevalence of 38.4% (Figure 3A; Table 2; p=0·03).

**Table 2:** Seropositivity by different characteristics of the study population

| Characteristics | Level | Seropositivity<br>Sample-weighted (95%CI) | p-value |
| --- | --- | --- | --- |
| **Subward** | Yerwada | 55.5 (46.6, 64.1) | <0.001 |
|  | Kasba Peth-Somwar Peth | 35.8 (29.2, 43.1) |  |
|  | Rasta Peth-Ravivar Peth | 44.1 (35.7, 52.8) |  |
|  | Lohiyanagar-Kasewadi | 66.4 (57.8, 74.1) |  |
|  | Navi Peth-Parvati | 54.1 (48.3, 61.7) |  |
| **Age categories**<br>**(years)** | 18-30 | 51.5 (39.2, 63.6) | 0.030 |
|  | 31-50 | 52.3 (41.0, 63.4) |  |
|  | 51-65 | 54.6 (39.8, 68.7) |  |
|  | 66-above | 38.4 (27.6, 50.6) |  |
| **Gender** | Male | 52.7 (41.7, 63.5) | 0.209 |
|  | Female | 49.7 (37.5, 62.0) |  |
| **High school education** | Completed | 42.2 (31.2, 54.1) | 0.013 |
|  | Not completed | 55.9 (44.3, 66.9) |  |
| **Type of dwelling** | Non-slum | 34.4 (26.2, 43.7) | 0.002 |
|  | Slum | 59.2 (48.8, 68.9) |  |
| **Shared toilet** | No | 45.3 (33.6, 57.5) | 0.008 |
|  | Yes | 61.7 (49.8, 72.4) |  |
| **Family size** | < 2 | 42.0 (31.6, 53.1) | 0.053 |
|  | 2 to 5 | 50.3 (39.3, 61.3) |  |
|  | > 5 | 58.7 (43.2, 72.6) |  |
| **Floor area per person** | < 5 m<sup>2</sup> | 63.4 (52.9, 72.8) | 0.002 |
|  | 5 to 9 m<sup>2</sup> | 52.7 (37.5, 67.4) |  |
|  | 10 and > m<sup>2</sup> | 35.8 (27.4, 44.1) |  |
| **Contact with case** | Yes | 55.4 (40.1, 69.8) | 0.371 |
|  | No | 51.0 (39.6, 62.2) |  |
| **History of symptom** | Yes | 62.5 (40.6, 80.2) | 0.088 |
|  | No | 50.5 (39.5, 61.4) |  |
| **History of COVID-19** | Yes | 90.5 (51.7, 98.9) | 0.030 |
|  | No | 50.7 (39.4, 62.0) |  |

**Figure 2:**
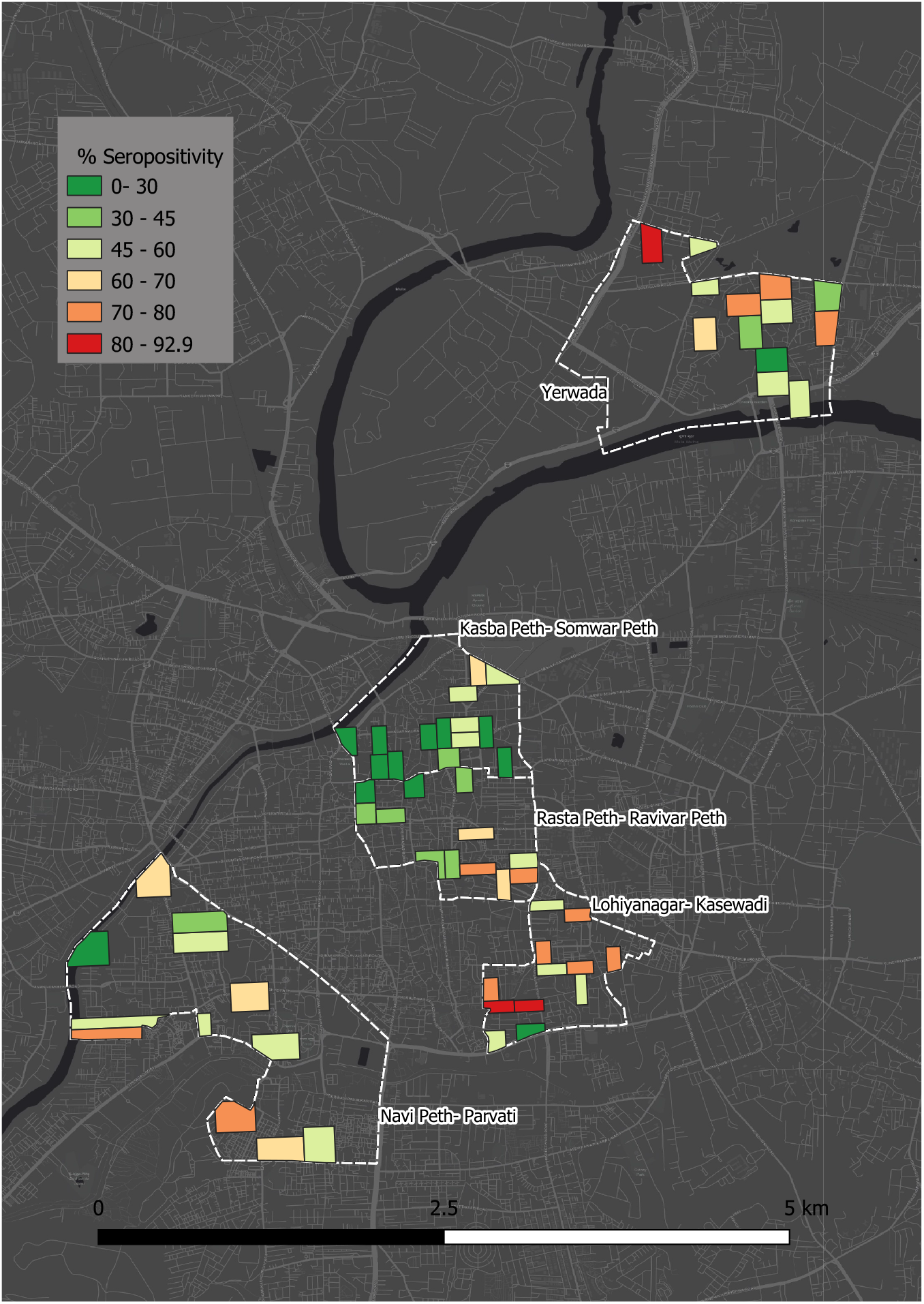
Map showing the selected subwards (dotted white outline) with the estimated seroprevalence in the sampled clusters. Military areas, prisons, educational institutions and riverbeds were excluded from the sampling.

**Figure 3:**
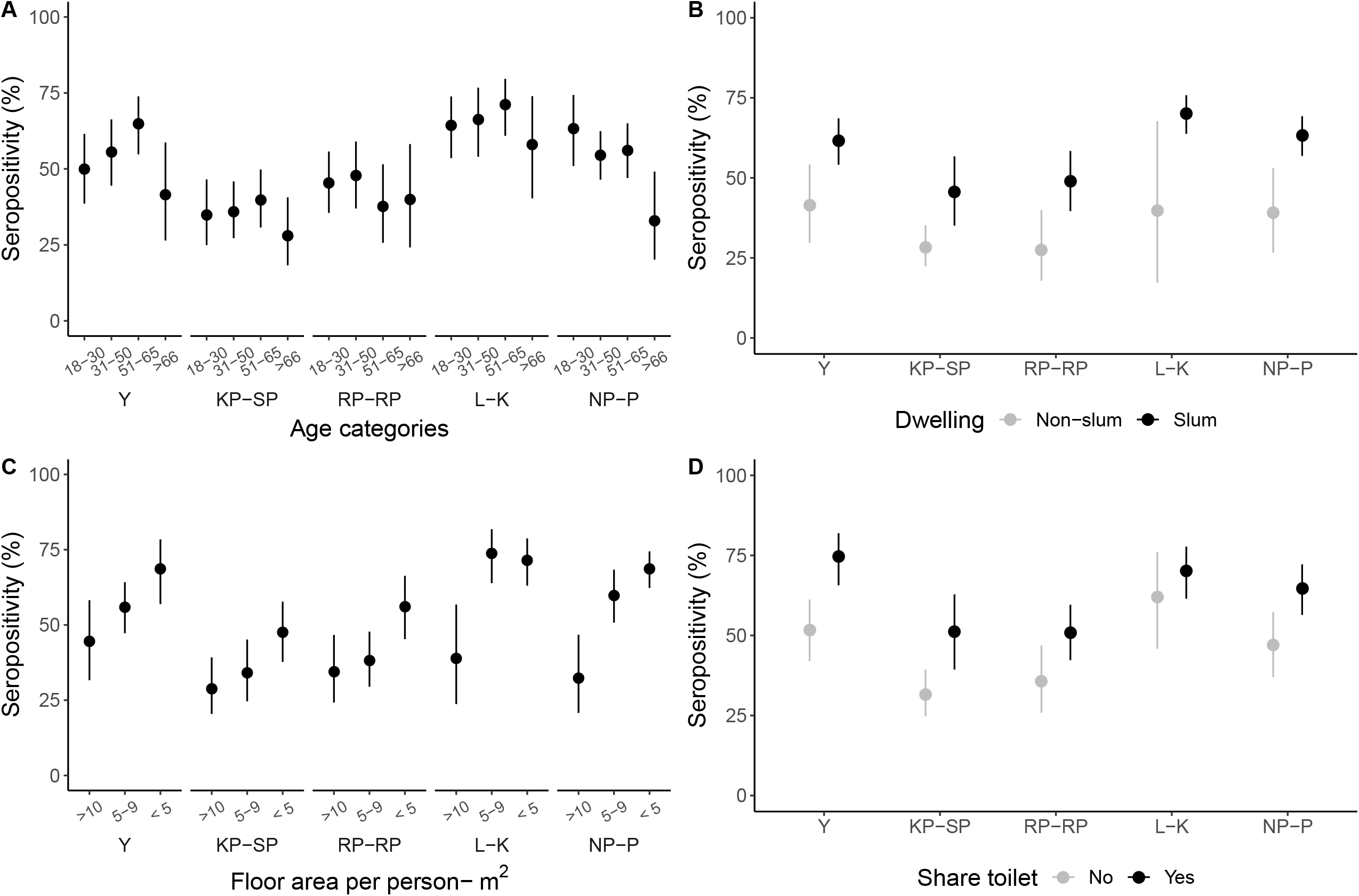
Association of seroprevalence (dot: mean prevalence; line: 95%CI) with age (**A**), type of dwelling (**B**), per capita floor area in m^2^ (**C**) and toilet use (**D**) in each subward.

Those living in densely populated slums had a higher seroprevalence (Table 2; Figure 3B; 59·2%; 95%CI 48·8 to 68·9) as compared to those living in non-slum areas (34·4%; 95%CI 26·6 to 43·7) and 63.4%(95%CI 52.9 to 72.8) of those living in crowded dwellings with < 5 m^2^ per capita floor space were seropositive compared to 35.8% (95%CI 27·4 to 44·1) for those with ≥ 10 m^2^ (Table 2; Figure 3C). Similarly, those using a shared toilet had a seroprevalence of 61·7% (95%CI 49·8 to 2·4) compared to 45·3% (95%CI 33·6 to 57·5) among those who did not (Table 2; Figure 3D). The presence of chronic illness, travel history, working status, exposure to COVID-19 patient, history of quarantine, participation in social gatherings, self/family members working in health care set up did not reveal any significant association with seropositivity.

Multiple logistic regression model revealed lower odds of SARS-CoV-2 seropositivity among those above 65 years of age (OR 0·59; 95%CI 0·36 to 0·98; *p*=0.044) as compared to those between 18-30 years (Table 3). Living in slums (OR 1·91; 95%CI 1·34 to 2·73; *p*=0.007) or in dwellings with per-capita floor space <5 m^2^ (OR 2·09; 95%CI 1·43 to 3·04) were also identified as independent risk factors for seropositivity (Table 3).

**Table 3:** Factors associated with seropositivity

|  | Unadjusted |  | Adjusted |  |
| --- | --- | --- | --- | --- |
|  | OR (95% CI) | p-value | OR (95%CI) | p-value |
| **Age category (years)** |  |  |  |  |
| 18-30 | 1 |  | 1 |  |
| 31-50 | 1.04 (0.76, 1.40) | 0.767 | 1.00 (0.71, 1.41) | 0.990 |
| 51-65 | 1.14 (0.72, 1.80) | 0.488 | 1.07 (0.61, 1.86) | 0.758 |
| >65 | **0.59 (0.35, 0.99)** | **0.047** | **0.59 (0.36, 0.98)** | **0.044** |
| **Gender** |  |  |  |  |
| Male | 1 |  | 1 |  |
| Female | 0.89 (0.71, 1.11) | 0.209 | 0.86 (0.69, 1.07) | 0.134 |
| **High school education** |  |  |  |  |
| Completed | 1 |  | 1 |  |
| Not completed | **1.74 (1.21, 2.49)** | **0.013** | 1.26 (0.92, 1.74) | 0.112 |
| **Type of dwelling** |  |  |  |  |
| Non-slums | 1 |  | 1 |  |
| Slums | **2.77 (1.88, 4.07)** | **0.002** | **1.91 (1.34, 2.73)** | **0.007** |
| **Family size** |  |  |  |  |
| < 2 | 1 |  |  |  |
| 2 to 5 | 1.40 (0.81, 2.41) | 0.159 |  |  |
| > 5 | 1.97 (0.94, 4.11) | 0.063 |  |  |
| **Floor area per person** |  |  |  |  |
| >10 m<sup>2</sup> | 1 |  | 1 |  |
| 5-9 m<sup>2</sup> | **2.00 (1.13, 3.54)** | **0.028** | 1.53 (0.92, 2.56) | 0.082 |
| < 5 m<sup>2</sup> | **3.12 (2.00, 4.86)** | **0.002** | **2.09 (1.43, 3.04)** | **0.004** |
| **Shared toilet** | 1.95 (1.33, 2.85) | 0.008 | 1.12 (0.83, 1.53) | 0.353 |
| **History of symptoms** | 1.63 (0.89, 3.01) | 0.090 | 1.54 (0.88, 2.69) | 0.099 |
| **History of COVID-19** | 9.30 (0.95, 91.11) | 0.053 |  |  |

The instantaneous reproduction number (R) declined following lockdown but remained largely above 1.0 (Figure 4 A-E). The reported incidence of COVID-19 after the serosurvey (between Aug 01 and Nov 23) was associated with seroprevalence in each subward (Slope=-0·74, *p*=0·007), after adjusting for the reported incidence at initiation of serosurvey (Figure 4 F).

**Figure 4:**
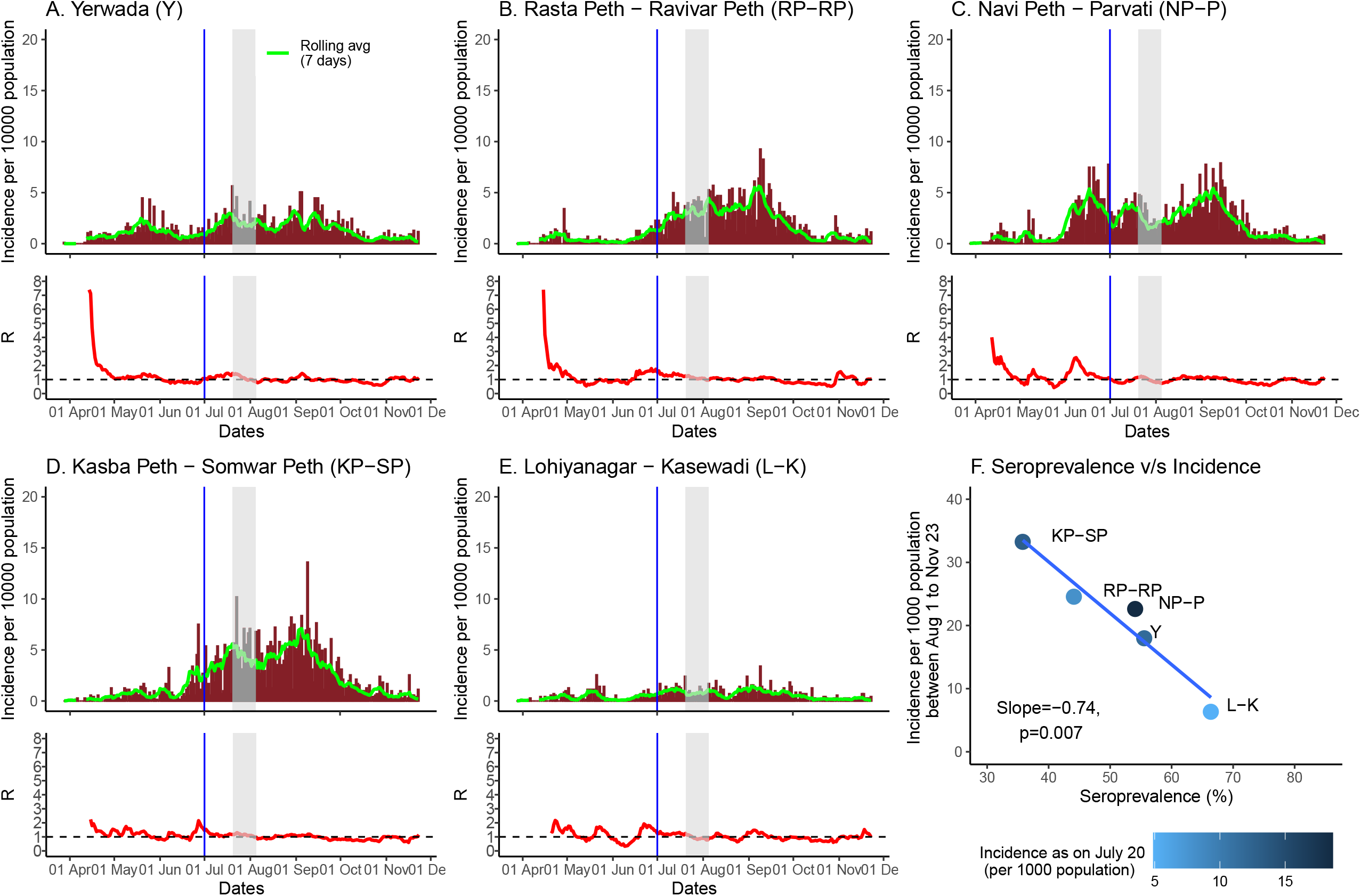
**A-E**, Time series of the reported SARS-CoV-2 infections (top) and the instantaneous reproductive number (R; bottom) in each subward. The light-grey region indicates the duration of sample collection and the blue line is the reference date for the serosurvey. **F**, The association between seroprevalence and new infections post serosurvey.

Using Government mortality data from the sampled sub-wards, we calculated an overall infection fatality ratio (IFR) of 0.21%, that was highest in Kasbapeth-Somwarpeth (0·28%) and lowest in Navipeth-Parvati (0·17%) (Figure 5). Older age-groups had higher IFRs compared to younger age-groups (Figure 5).

**Figure 5:**
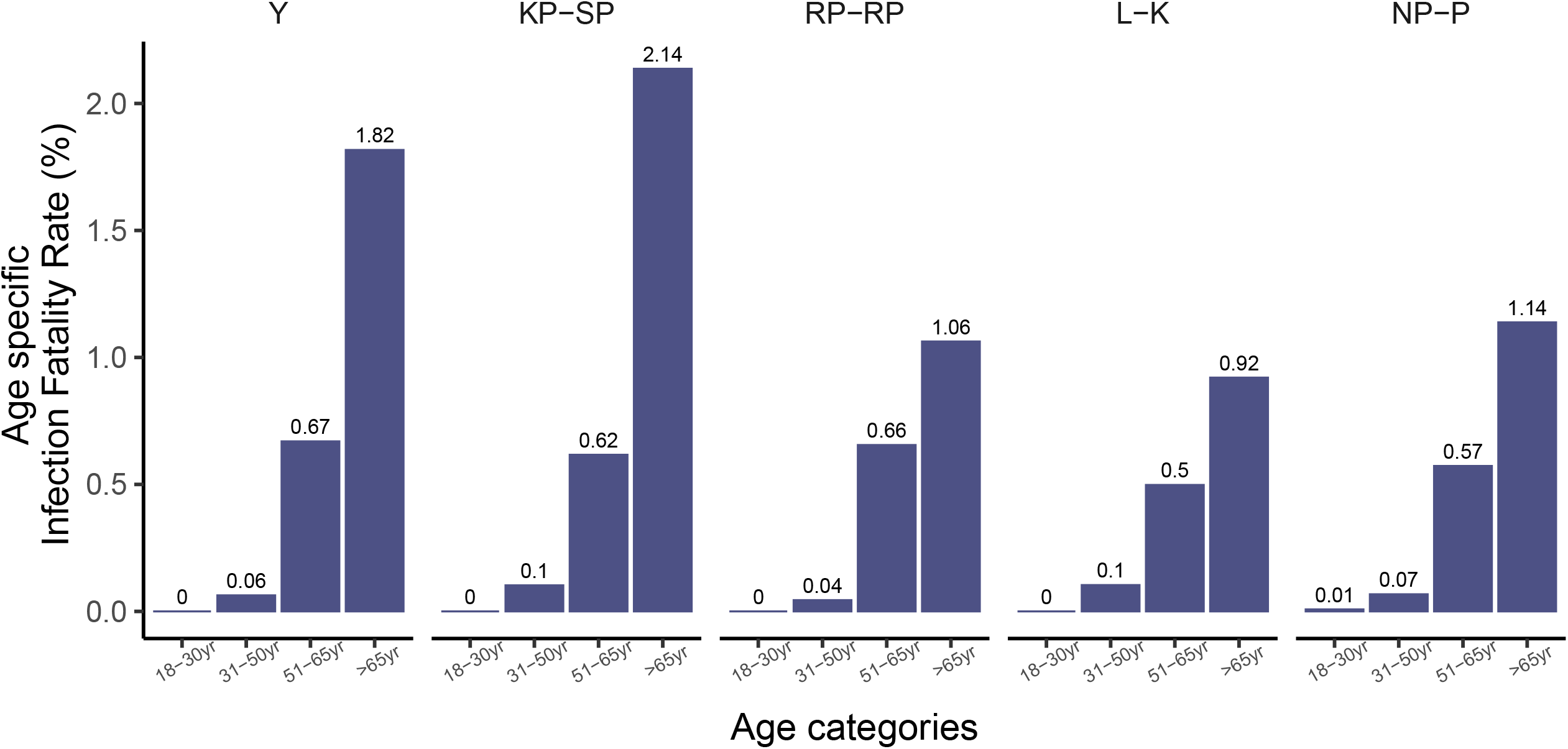
Estimated Infection Fatality Rates (IFR) in each subward stratified by age.

### Receptor-binding inhibition activity in the asymptomatic population

From the 857 RBD-IgG positive sera, 697 samples were randomly selected and tested for the ability to inhibit the binding of SARS-CoV-2 RBD to human ACE-2 using a specific ELISA-based surrogate virus neutralization test (sVNT)^20^. 85·1% of the RBD-IgG positive sera were sVNT positive.

While 57·2% of 18-30 years age-group showed ≥50% inhibition, this increased to 64% in the 31-50 years category and became 75% in the population above 50 years (*p*<0.001). However, no association was found with sex.

An explanation for the age-association could be the association of RBD-binding antibody levels with age, as high sVNT values are likely to be positively correlated with high RBD-binding activity. The RBD-IgG ELISA signal to cut-off ratio (S/Co) was indeed positively correlated with the % inhibition of RBD-ACE-2 binding (Figure 6A; R = 0·718; 95%CI 0·634 to 0·717; *p*<0·0001) and higher levels of surrogate neutralisation was observed in RBD-IgG high S/Co (≥3) samples compared to low RBD-IgG (Figure 6B; p<0.0001). However, the RBD-IgG levels were not associated with age, making the sVNT association with age mechanistically intriguing.

**Figure 6:**
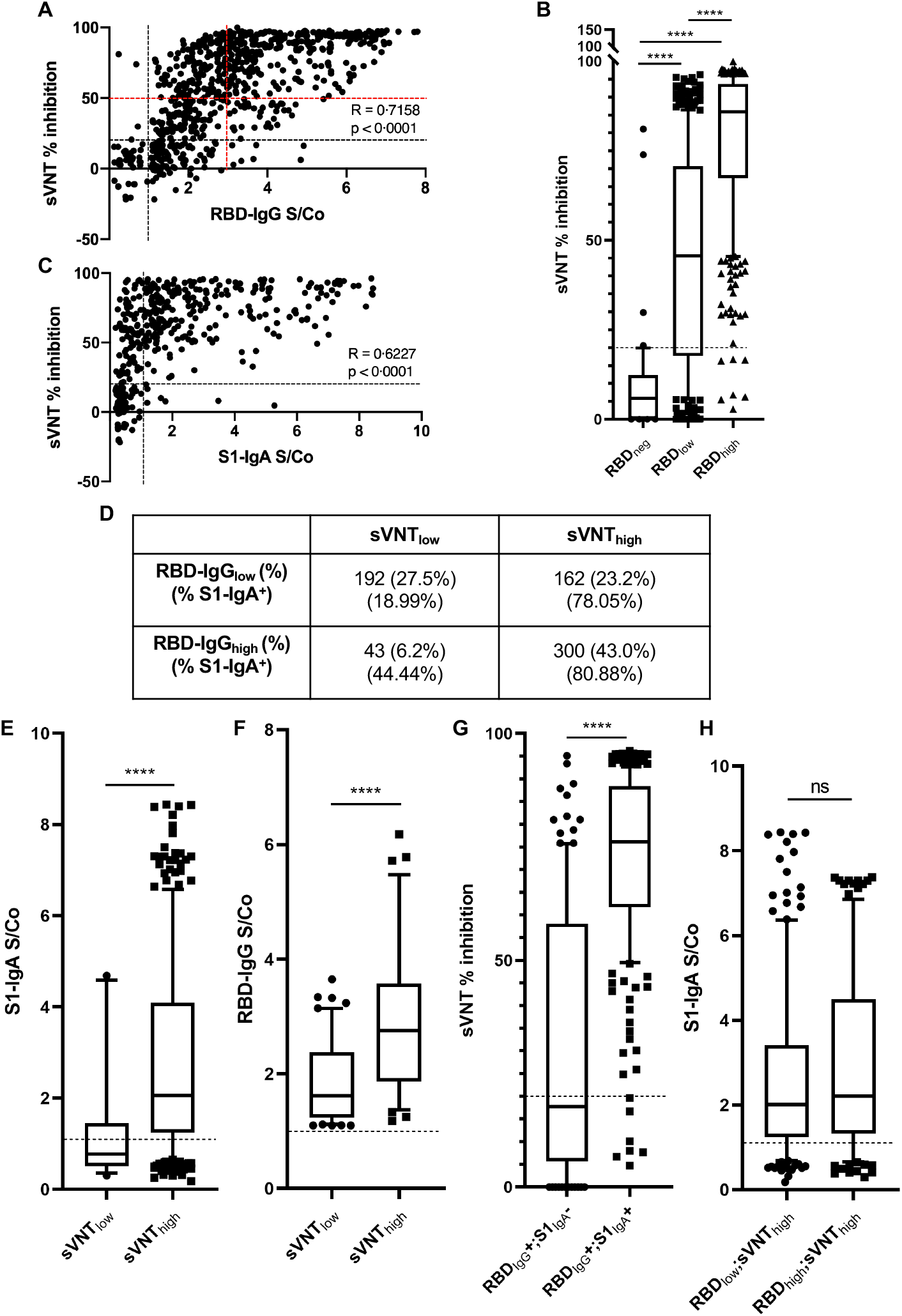
**A**, Correlation between percent inhibition in the sVNT and RBD-IgG ELISA S/Co ratio (r=0·7158; *p*<0·0001). The red dashed lines indicate an arbitrary cut-off for high/low values for both assays (RBD-IgG ≥3; sVNT % inhibition ≥50). **B**, Comparison of sVNT % inhibition between RBD-IgG levels. **C**, Correlation between percent inhibition in the sVNT and S1-IgA ELISA S/Co ratio (r=0.6227; *p*<0.0001). **D**, Head-to-head analysis of high/low sVNT % inhibition with high/low RBD-IgG categories and associated S1-IgA positivity (within each category). **E-F**, Comparison of S1-IgA ELISA and RBD-IgG S/Co ratios among high/low sVNT % inhibition. **G**, Comparison of sVNT % inhibition between S1-IgA positive and negative groups among RBD-IgG positive cases. **H**, Association of S1-IgA levels with low/high RBD-IgG showing high sVNT % inhibition. The black dashed lines indicate developer/manufacturer defined cut-off values. ****, *p*<0.0001; ns, non-significant.

To explore the heterogeneity in the association between RBD-IgG and surrogate neutralisation (Figure 6A,D), we classified the response into four categories (across high/low RBD-IgG and high/low surrogate neutralization) using arbitrary cutoffs of 3 (RBD-IgG S/Co) and 50% (surrogate neutralization). We found no correlation with either age or sex in these four categories. The heterogeneity in the response was interesting in two distinct ways. The high RBD-IgG/low-inhibition group (6.2%) indicates that a significant subpopulation may not be well protected despite having high RBD-IgG. The presence of a large high-inhibition/low RBD-IgG sub-population (23.2%) was also intriguing.

A possible explanation for the low RBD-IgG/high-inhibition group were non-IgG RBD-binding antibodies. We therefore tested a subset of 639 samples for IgA against the S1 domain of the SARS-CoV-2 spike protein. 68·4% of the RBD-IgG positive sera were also S1-IgA positive. Of the RBD-IgG negative sera, only 8·6% had detectable S1-IgA. S1-IgA S/Co was positively correlated with the surrogate neutralisation (Figure 6C; R=0·623; 95%CI 0·557 to 0·681; *p*<0·0001). 79·21% of samples with ≥50% inhibition in the sVNT were S1-IgA positive (Figure 6D). Both S1-IgA as well as RBD-IgG levels were higher in the high sVNT category (Figure 6E, F). The low RBD-IgG/high-inhibition category also had a high proportion of S1-IgA positivity (78·05%) compared to low RBD-low inhibition category (18·99%) (Figure 6D). RBD-IgG and S1-IgA double-positive sera were associated with significantly higher surrogate neutralisation than RBD-IgG single positive (Figure 6G; *p*<0.0001). However, neither the frequencies of S1-IgA positivity nor the S1-IgA S/Co values were different between the low versus high RBD-IgG subgroups within the high inhibition category (Figure 6D,H), suggesting the possibility of a more complex biological basis for the low RBD-IgG/high-inhibition subgroup.

In summary, there is significant surrogate neutralisation in the sera of individuals asymptomatically infected with SARS-CoV-2 with likely contributions from both IgG and IgA antibodies. However, this functional response shows considerable heterogeneity, with patterns of variation suggesting age-associated diversity in the antibody repertoire.

### Virus (pandemic strain) neutralising activity correlates with RBD-IgG and surrogate neutralisation

To estimate the association of RBD-IgG with neutralising activity, we evaluated neutralising activity in a subset of sampled sera (n=192) by estimating PRNT_50_ values. Compared to RBD-IgG negative sera (median titre_neg_=0), both low and high levels of RBD-IgG positive sera had significantly higher neutralising antibody titres (Figure 7A; median titre_≤3_=80, IQR 20-320, *p*_*≤3*_<0·0001; median titre_≥3_=320, IQR 160-1280, *p*_*≥3*_<0·0001).

**Figure 7:**
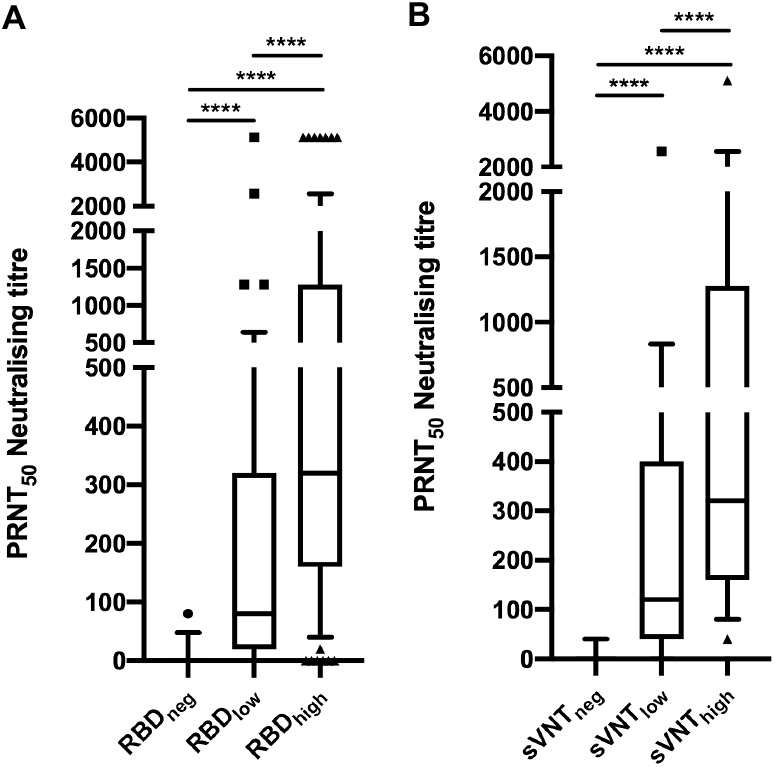
Comparison of *in vitro* virus neutralisation activity across RBD-IgG (**A**) and sVNT inhibition (**B**) levels. ****, *p*<0·0001.

Similarly, compared to sVNT negative sera (median titre_neg_=0), substantial virus neutralising titres were observed in sera with both low and high inhibition in the sVNT (Figure 7B; median titre_≤50%_=120, IQR 40-400, *p*_*≤50%*_<0·0001; median titre_≥50%_=320, IQR 160-1280, *p*_*≥50%*_<0·0001).

These results indicate a strong association of both RBD-IgG levels and surrogate neutralisation with live virus neutralization activity *in vitro*.

### Systemic inflammation in asymptomatic SARS-CoV-2 infections

We evaluated C-reactive protein (CRP), a marker for systemic inflammation^21^, levels in all the samples collected. No gross differences were found in CRP levels between the RBD-IgG positive and negative sera. Thus, there was no evidence of residual systemic inflammation in asymptomatic SARS-CoV-2 infections. In both groups, women and those aged over 50 years had higher CRP levels (≥3000 ng/ml; data not shown). Interestingly, sera with high levels of surrogate neutralisation (≥ 50%) also showed significantly higher CRP levels (Figure 8; *p*=0.0014). It may be recalled here that high sVNT inhibition levels were associated with older age groups that also have higher CRP levels. However, high RBD-IgG levels were not associated with high CRP levels, just as they were not associated with age, making the age and CRP association with surrogate neutralisation of substantive biological interest.

**Figure 8:**
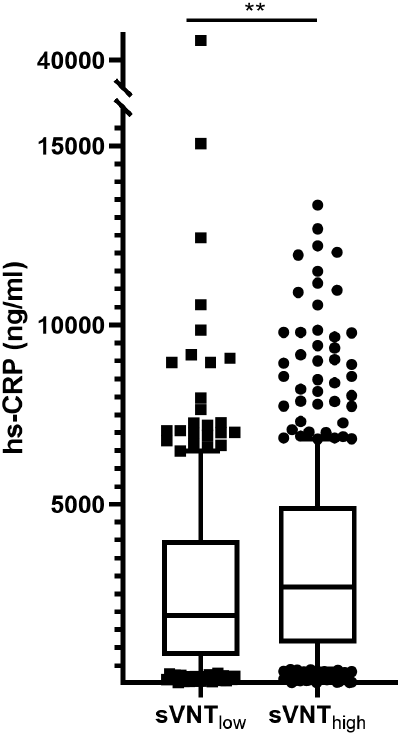
Comparison of hs-CRP (ng/ml) levels among low/high sVNT % inhibition categories. **, *p*=0·0014.

## DISCUSSION

This is the first systematic study (at the time of submission) from a LMIC reporting community SARS-CoV-2 sero-surveillance of high granularity alongside estimation of correlates of immune protection. We estimated seroprevalence as well as serological correlates of protection in a cross-sectional cohort of 1659 asymptomatic participants from five small urban localities in the metropolitan city of Pune, India. IgG seroprevalence was determined against the receptor-binding-domain (RBD) of the SARS-CoV-2 spike protein, to aid correlation with immune protection since RBD is the predominant target for neutralising antibodies. Large subsets of the sera were also tested for surrogate neutralisation as well as live SARS-CoV-2 virus neutralisation, data not so far reported in community sero-surveillance studies.

Analysis of anti-RBD IgG seroprevalence revealed extensive SARS-CoV-2 infection in the sampled population. Despite the proximity of the sampled subwards within Pune city, there was variation across these locations suggesting that local factors influence prevalence. Across the subwards, the seroprevalence was higher amongst slum dwellers compared to those living in apartments and bungalows. Residential crowding and shared sanitation were associated with increased seroprevalence. Collectively, these observations suggest high intrafamilial transmission, though our sampling strategy precluded direct analysis.

While infection risk was similar between men and women, the seroprevalence in the elderly population (65+ age-group) was lower than the younger population. This observation is in contrast to what has been reported from Delhi^22^ and Mumbai^3^ but concordant with trends reported from meta-analysis of global studies^1^. Our observation might reflect limited mobility and higher compliance with preventive measures in this age-group. A lower IgG response to infection and/or accelerated seroreversion in the older population is also possible. However, the latter may not be likely as no association with age was found for either RBD-IgG or S1-IgA levels.

We estimated an overall IFR of 0.21% based on reported fatalities. This is consistent with a global median IFR of 0.27%, estimated from a meta-analysis of multiple studies^23^. The IFR ranged from 0.17% to 0.28% across the subwards and in line with reports from multiple studies world-wide^23^, the IFR increased with age.

The substantial locality-specific variations in seropositivity levels and infection fatality rates (IFRs) highlight heterogeneities of infection behaviour even in dense, urban populations often lost in more global analyses.

An inverse correlation was observed between subward-level seroprevalence and the number of positive cases reported in the subsequent period which persisted after adjusting for baseline disease incidence rates to account for any biased testing. This observation revealed a strong negative association with seropositivity, indicating potential modification of transmission by community immunity. This also suggests population-level protection, at least in the short-term when seroprevalence approaches herd immunity threshold. These trends should be interpreted with caution given our dependence on publicly available data and limited understanding of the durability of immune responses.

We used RBD-IgG for our analysis of seroprevalence. Compared to nucleocapsid IgGs, anti-RBD/Spike IgGs are strongly correlated with virus neutralising activity, have higher sensitivity, and display comparatively less cross-reactivity to other coronaviruses^4 5 24^. We leveraged this to directly test for virus neutralising activity by estimating the inhibition of Spike protein binding to the ACE-2 receptor. 85.1% of the asymptomatic population showed surrogate neutralisation. RBD-IgG and sVNT positivity were both correlated with significant neutralisation titres in PRNTs. Thus elevated RBD-IgG and surrogate neutralisation are robust predictors of virus neutralising activity.

The RBD-IgG S/Co was strongly correlated with surrogate neutralisation, though significant heterogeneity was observed around this response. Approximately 30% of individuals showed significant departures from this correlation, underlining significant immune response heterogeneities. This heterogeneity may be of biological and clinical relevance but we found no association with either age or sex. Non-IgG antibody isotypes were speculated to contribute to the low RBD-IgG/high inhibition category. Since the IgM response is relatively more transient^4^, IgA was evaluated. However, neither the proportion or levels of IgA differed significantly between the low RBD-IgG/high inhibition and high RBD-IgG/high-inhibition categories. Therefore, it is plausible that individuals in this category may have developed functionally different antibody repertoires. This could consist of a greater prominence of neutralising epitopes or antibodies with greater affinity, since inhibition/neutralisation activity would be more sensitive to affinity than binding activity. Interestingly, the older population showed high levels of surrogate neutralisation more frequently than the young, yet there was no age association with the IgG and IgA response. This could imply a shift in the epitope dominance towards neutralizing epitopes, and/or higher affinities in the older population.

68·4% of RBD-IgG positive individuals were also S1-IgA positive. As anti-RBD IgA is short-lived in comparison to anti-RBD IgG^4^, the double-positive population may represent more recent infections. Given the higher surrogate neutralisation activity in the double-positive individuals, it could be speculated that immune protection may wane over time. This has implications on population-level protection from infection.

Unlike in symptomatic cases^7-10^, there was no difference in CRP levels between the seropositive and seronegative populations indicating the absence of chronic elevation of systemic inflammation in asymptomatic infections. The association of higher CRP levels with higher inhibition levels, without a corresponding association with RBD-IgG or IgA levels, is more likely to reflect the age correlation of high inhibition levels, rather than any specific association between persistent inflammation and high inhibition activity.

Our study had several limitations. We focussed on the seroprevalence in settings reporting the highest incidence, in part due to concerns about the impact of imperfect diagnostic accuracy on seroprevalence in low prevalence settings. This along with the limited sample size made it challenging to extrapolate seroprevalence more broadly. For operational reasons, we excluded active containment zones, those with an acute illness and restricted sampling to one individual per household; these could have biased the seroprevalence estimates. Our study did not include children who may have an important role in transmission.

Nonetheless, this study, demonstrates the heterogenous, but widespread transmission of SARS-CoV-2 in a dense urban population. The high seroprevalence combined with evidence of neutralizing antibodies and of declining incidence in settings of high seroprevalence raises the possibility of population immunity, at least in the short term.

That indoor crowding, living in a slum and sharing a common toilet were significant predictors of seropositivity highlights the challenges India faced in controlling the pandemic despite stringent lockdown measures. Despite this, the relative protection of the elderly, and low infection fatality ratios and evidence of population immunity provide hope that if control measures continue, urban India may well have passed the peak of the pandemic. High seroprevalence in the dense urban localities of the study site, despite a protracted and stringent lockdown, provides a realistic account on transmission dynamics crucial for public health policies in LMICs. Micro-geographic variability within locales, dominated by sub-optimal living conditions, needs to be acknowledged and used to develop measures designed for people in such socio-economic contexts. As a next step, further assessment of the immunity and infection rates in settings of high seroprevalence in other majorly affected cities are needed. The heterogeneity of correlation between RBD seropositivity and neutralising capacity, as well as the complex association with age encountered in this study open up a plethora of research questions into epitope dominance and affinity variations in anti-viral antibodies in asymptomatic infection.

## Data Availability

All data is available on reasonable request.

## AUTHOR CONTRIBUTIONS

AG* contributed in study design, study coordination, funding acquisition, analysis, interpretation and manuscript writing; SB contributed in data acquisition; ASK contributed in analysis and writing manuscript; AK contributed in field coordination, sample and data acquisition; JMM data acquisition/curation; AJ contributed in data acquisition; GM contributed in data acquisition; GK contributed in study design, interpretation and writing manuscript; VB contributed in study design, interpretation, writing manuscript; SR contributed in study design, interpretation and writing manuscript; LSS* contributed in study design, study coordination, funding acquisition, analysis, interpretation and writing manuscript; JJ* contributed in study design, analysis, interpretation and manuscript writing; SC* contributed in study design, data acquisition, analysis, interpretation and writing manuscript; AN* contributed in study design, study coordination, analysis, interpretation and writing manuscript.

*Joint senior authors

## TRANSPARENCY DECLARATION

The corresponding author* affirms that this manuscript is an honest, accurate, and transparent account of the study being reported; that no important aspects of the study have been omitted; and that any discrepancies from the study as planned (and, if relevant, registered) have been explained.

*The manuscript’s guarantor.

## PATIENT AND PUBLIC INVOLVEMENT

Patients or the public were not involved in the design, or conduct, or reporting, or dissemination plans of our research

## DISSEMINATION DECLARATION

Apart from uploading the preprint in a publicly accessible preprint server, to targeted dissemination is applicable.

## DATA SHARING

All anonymised data are available for sharing upon reasonable request

## ACKNOWLEDGEMENTS

The Persistent Foundation is acknowledged for financial support. We thank the Pune Municipal Corporation, legislators, commissioners, and medical officers for facilitating sample collection. All members of the sample collection and processing teams are acknowledged. We also acknowledge administrative and infrastructural support from IISER Pune, SPPU, THSTI, and CMC Vellore. We acknowledge Prof. Raghavan Varadarajan, IISc Bengaluru for his kind gift of the RBD expression construct. All non-author contributions are listed in the Supplementary file.

