## Supplementary Information for "Community prevalence of antibodies to SARS-CoV-2 and correlates of protective immunity in an Indian metropolitan city"

**Acknowledgement of non-author contributions**

The authors acknowledge the following individuals for technical support.

| Name | Affiliation | Role |
|---|---|---|
| Kajal Medipilwar | SPPU Pune | Sample collection |
| Sanika More | SPPU Pune | Sample collection |
| Majed Hussain | SPPU Pune | Sample collection |
| Snehal Kulkarni | SPPU Pune | Field data management and analysis |
| Amruta Kulkarni | SPPU Pune | Field data management and analysis |
| Ketakee Ghate | IISER Pune | Sample processing |
| Sanket Nagarkar | IISER Pune | Sample processing |
| Dhriti Nagar | IISER Pune | Sample processing |
| Devika Bodas | IISER Pune | Sample processing |
| Pooja Vaid | IISER Pune | Sample processing |
| Madhura Kulkarni | Prashanti Cancer Care Mission Pune | Sample processing |
| Devdatta Tengshe | GeoSpoc Geospatial Services Pvt. Ltd. | Geospatial mapping |
| Rahul Chopra | Independent researcher, formerly IISER Pune. | Geospatial mapping |
| Subhash Tanwar | THSTI Faridabad | Serological assays |
| Manisha Yadav | THSTI Faridabad | Serological assays |
| Simran Babbar | THSTI Faridabad | Serological assays |
| Himakshi Sidhar | THSTI Faridabad | Serological assays |
| Imran Khan | THSTI Faridabad | Serological assays |
| Chanchal Sharma | THSTI Faridabad | Serological assays |
| Sudipta Deepak Sonar | THSTI Faridabad | Serological assays |

| | | |
|---|---|---|
| Kamini Jakhar | THSTI Faridabad | Serological assays |
| Mahima Tiwari | THSTI Faridabad | Resource support for serological assays |
| Deepak Chaurasiya | THSTI Faridabad | Resource support for serological assays |
| Shailendra Mani | THSTI Faridabad | Resource support for serological assays |
| Tripti Srivastava | THSTI Faridabad | Resource support for serological assays |
| Sneha Bhogale | Pune Knowledge Cluster Pune | Public data compilation |
| Ashwini Keskar | Pune Knowledge Cluster Pune | Public data compilation |
| Bhalchandra Pujari | SPPU Pune | Public data analysis |
| Sarika Bhattacharya | NCL Pune | Public data analysis |
| Anu Raghunathan | NCL Pune | Public data analysis |
| Shilpa Jain | IISER Pune | Administrative management |
